# Framework for enhancing the estimation of model parameters for data with a high level of uncertainty

**DOI:** 10.1101/2020.12.17.20248389

**Authors:** Gustavo B. Libotte, Lucas dos Anjos, Regina C. Almeida, Sandra M. C. Malta, Renato S. Silva

**Author notes:** E-mail: {, }.

## Abstract

Reliable data is essential to obtain adequate simulations for forecasting the dynamics of epidemics. In this context, several political, economic, and social factors may cause inconsistencies in the reported data, which reflect the capacity for realistic simulations and predictions. In the case of COVID-19, for example, such uncertainties are mainly motivated by large-scale underreporting of cases due to reduced testing capacity in some locations. In order to mitigate the effects of noise in the data used to estimate parameters of models, we propose strategies capable of improving the ability to predict the spread of the diseases. Using a compartmental model in a COVID-19 study case, we show that the regularization of data by means of Gaussian Process Regression can reduce the variability of successive forecasts, improving predictive ability. We also present the advantages of adopting parameters of compartmental models that vary over time, in detriment to the usual approach with constant values.

## 1 Introduction

Since the onset of the novel coronavirus (SARS-CoV-2) pandemic, at the end of 2019, a wealth of research has been carried out from across the globe, aiming to understand the dynamics and transmission patterns of the disease. More than a year after the notification of the first case, the number of infected individuals keeps rising significantly worldwide. In the meantime, the confirmation of reinfections and identification of seasonal immunity [14] reinforces the need for actions to contain the disease, even in locations where the epidemic would be under control.

Governmental decisions to mitigate the spread of the disease, such as the introduction of lockdown and social distancing measures, are usually based on computational simulations whose preeminent objective is to predict the way the disease spreads in the population, considering continuously reported data [12]. However, there are several factors associated with natural, economic, and social aspects that make it difficult to adequately predict the spread of the disease and, consequently, the definition of a comprehensive policy for prevention and control of the disease [40, 16, 31]. The heterogeneity of the population in relation to attributes such as demographic diversity, age-dependent characteristics, and randomness related to the mobility and interaction of individuals makes it hardly possible to create a model capable of incorporating all these features together (see, for instance, Refs. [4, 7, 9, 13]).

Asymptomatic people also play a significant role in the ongoing pandemic. Oran and Topol [30] presented a comprehensive bibliographic review on the estimation of asymptomatic cases of COVID-19 in different parts of the world and concluded that the proportion of asymptomatic individuals may vary from 40% to 45% in relation to the total number of reported cases. The scenario of widespread underreporting coupled with a deficient screening and testing capacity leads to significant uncertainty in relation to the reported data of infected individuals. The impact of such uncertainties was analyzed by Ioannidis [17], Li et al. [21], and Wu et al. [51].

Turning attention to the impact of the pandemic in Brazil, a country with a continental dimension, such problems tend to become even more evident [23]. Socioeconomic inequalities [37, 48] and cultural factors [11, 33] have a direct impact on access to information and health services, which translates into high rates of infection and, as a consequence, underreporting cases. Veiga e Silva et al. [49] also analyzed presumptive inconsistencies in the data collected and made available by the Ministry of Health in Brazil. They reported that there may be a difference of approximately 41% in the number of deaths caused by COVID-10 related complications.

In an attempt to describe the spread of COVID- 19 on different population groups, several models have been proposed by means of integrating typical features related to the disease, such as the quarantine period, lockdown, social distancing, and hospitalization. Massonis et al. [24] bring together several of these models, classifying them hierarchically in relation to the number of coupled features. In general, even the most complex models, that is, those with supposedly more capacity to associate knowledge about the spreading dynamics of the disease, tend to experience some difficulties in identifying the behavior of noisy data in the long-term, as shown by Alberti and Faranda [1] and Roda et al. [40].

The major motivation of this work is to provide alternatives to enhance the capacity of estimating parameters in compartmental models and predicting the spread of COVID-19, taking into account data with a high level of uncertainty, such as the number of new cases reported in the state of Rio de Janeiro since March 05, 2020. The data do not have well-defined behavior, in such a way that the variations in subsequent days are caused, in part, by the factors that deepen the disparities in the notifications of cases of infection. In addition, the Brazilian government estimates the COVID-19 data considering the daily count in the municipalities—Brazil is made up of 5570 municipalities, of which 92 make up the state of Rio de Janeiro—which are autonomous in relation to population testing policy and do not follow a common strategy to prevent the disease. As in some locations data are not reported on weekends, there are sudden drops in the number of new infections, which afterward cause unexpected increases when data are reported at the beginning of the following week.

In this context, aiming at expanding the predictive capacity of compartmental models subject to scenarios of great uncertainty, the objectives of this work are to propose the use of strategies capable of reducing the variability of the estimation of model parameters and to discuss the advantages of considering time-dependent parameters. We propose that the noisy data set be regularized by means of Gaussian Process Regression (GPR). This approach allows a set of data to be smoothed, so as to decrease its noise level without significantly changing its behavior. To confirm this assumption, we compared a subgroup of reported data on dead and infected individuals in the state of Rio de Janeiro to simulations produced by the SEIRPD-Q model, whose parameters are estimated using regularized data. The results obtained are also compared to the simulations calculated in the usual way, without regularizing the data, in order to quantify the predictive capacity in relation to known data and the gain in relation to the parameter estimation approach with unchanged data.

A recent review of the literature on the subject found that, to date, few works that incorporate the concepts of Gaussian Process (GP) applied to the epidemiological modeling of COVID-19 have been published. These works are briefly presented below. Ketu and Mishra [19] proposed the Multi-Task GPR model, aiming to predict the COVID-19 outbreak worldwide. The authors compared the results obtained with other regression models, in order to analyze the effectiveness of the proposed method. Zhou and Ji [52] proposed a model for transmission dynamics of COVID-19 considering underreporting of cases (what they called undocumented infections) and estimated the time-varying disease rate of transmission using GPR and a Bayesian approach. Arias Velásquez and Mejía Lara [2] demonstrated the correlation between industrial air pollution and infections by COVID-19 before and after the quarantine in Peru, by presenting a classification model using Reduced-Space GPR. This methodology is used by the same authors in Ref. [2] to report a long-term forecast for COVID-19 in the USA. In turn, Ribeiro et al. [38] compared the predictive capacity of various machine learning regression and statistical models, considering short-term forecasting of COVID-19 cumulative cases in Brazil. As far as we are aware, this is the first time that GPR is employed for regularization of COVID-19 data, which are subsequently analyzed using a compartmental model (although other methods for regularization have been applied [21]).

In our analysis, we partition the data sets between training and test data, and carry out successive parameter estimations varying the proportion between these types of data, in order to show the behavior of the obtained parameter set. We show that some of these parameters can be approximated by functions and, considering this possibility, we analyze the influence of adopting variable parameters over time. We use both a deterministic approach, in terms of least squares, to estimate the gain related to the use of regularized data and time-varying parameters, and a Bayesian approach, in order to analyze parameter uncertainties, which are propagated to the model to quantify uncertainties on the model outcomes.

## 2 Materials and methods

### 2.1 Compartmental model description

Following the models proposed by Jia et al. [18] and Volpatto et al. [50], we develop an extension of the Susceptible-Exposed-Infected-Removed (SEIR) model, termed SEIRPD-Q model, which further considers Positively Diagnosed (*P*) and Dead (*D*) groups of individuals. To account for social distancing measures, we implicitly consider a mechanism that isolates individuals from the virus transmission (–*Q*) rather than defining a specific compartment for individuals in quarantine. A schematic description of the SEIRPD-Q model is presented in Fig. 1. We consider a population susceptible to a viral outbreak, whose rate of transmission per contact is given by *β*. At the beginning of a COVID-19 infection, infected individuals pass through a latency period in which they are not capable of transmitting the disease to another individual, becoming infectious only after this stage, even without showing any sign of disease. Individuals in such conditions are said to be exposed, with incubation period given by 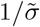. The infectious individuals are then divided into infected and positively diagnosed compartments, based on the premise that a large number of individuals who contract the virus are not diagnosed. This is due to the reduced testing capacity in some locations so that the diagnosis is made, primarily, in individuals who have severe symptoms or are hospitalized. The proportion of infected individuals, given by *ρ*, is related to the majority of people that only suffer mild symptoms and get recovered without significant complications. On the other hand, the complement of this group are those individuals who, in fact, have been positively diagnosed. In turn, individuals who recover from the disease after being in the infected compartment are moved to the removed compartment at a rate of *γ*_*I*_. The same goes for positively diagnosed individuals, who are removed at a rate of *γ*_*P*_. In addition, it is reasonable to assume that most of the individuals who died from complications caused by the disease had severe symptoms and were tested or hospitalized. Therefore, we do not consider that individuals in the infected compartment die from virus-related causes without being diagnosed, and the mortality rate of positively diagnosed individuals is given by *d*_*P*_.

**Fig. 1.**
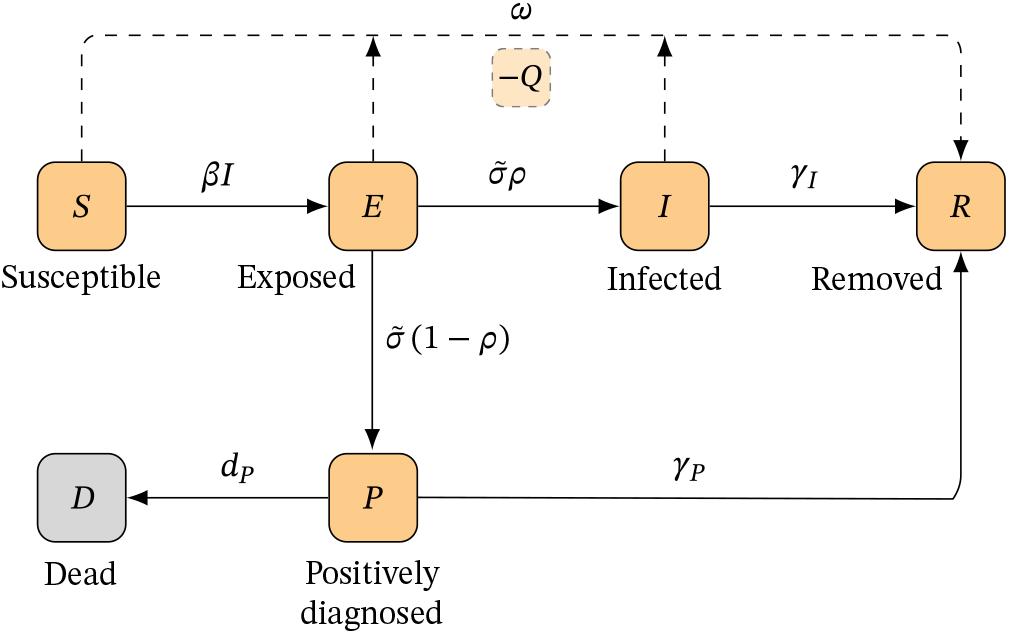
Schematic description of the SEIRPD-Q model.

Quarantine measures are also incorporated in this model, affecting the susceptible, exposed, and infected compartments. Individuals in these compartments are kept in quarantine at a rate of *ω*, and are not assumed to be infectious considering restrictive quarantine measures. The quarantine compartment is implicitly modeled and, therefore, the removed compartment includes individuals who have undergone quarantine, along with those who have been infected and have recovered from the disease. The formulation of the SEIRPD-Q model is given by the following system of ordinary differential equations:

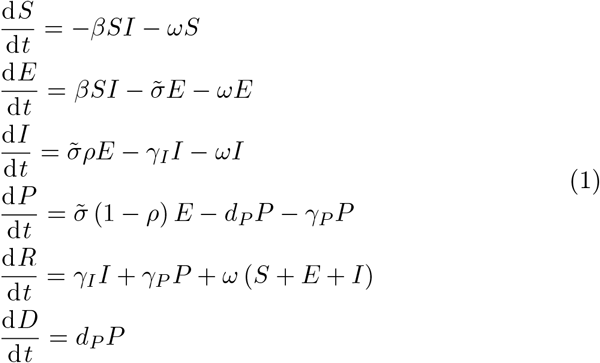

The SEIRPD-Q model presents some fundamental differences in relation to those on which it was based: first, Jia et al. [18] consider that only susceptible individuals are subject to quarantine measures, which is modeled using an explicit compartment and also considering an additional parameter that controls the social distancing relaxation; second, we disregard the asymptomatic compartment, according to Volpatto et al. [50], due to the lack of this information associated with limited testing of the population; third, we consider only the mortality rate of positively diagnosed individuals, unlike Volpatto et al. [50].

### 2.2 Data

The data used in this work are the daily number of infected (positively diagnosed) and dead individuals in the state of Rio de Janeiro. The Brazilian Ministry of Health reports the data daily, which are synthesized and made available as shown in Ref. [10]. The analyzed data refer to the period between March 10, 2020 and October 5, 2020, consisting of 210 records. Although the number of recovered individuals is also available, it may be wise not to use this data to estimate the parameters of the model since, due to the unseemly policy of testing the population, there is much uncertainty about these data.

In the following, the observable quantities are denoted by the time series 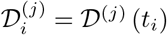, for *i* = 1, …, *p* and *j* ∈ {*P, D*}. Overall, let ***𝒟*** = {*𝒟*_1_, …, *𝒟*_*p*_} be the finite set of real-valued measurements collected from successive observations of the daily number of positively diagnosed and dead individuals at different times *t*_*i*_.

### 2.3 Deterministic approach for parameter estimation

Here we want to solve the so-called *inverse problem*, i.e., to determine the model parameters that generate outputs as close as possible to the observable data. To this end, let θ be the *m*-dimensional vector of model parameters, and 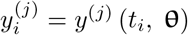 be the model responses at different times *t*_*i*_, *i* = 1, …, *p* and *j* ∈ {*P, D*}, as used previously. Additionally, let 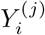 denote the cumulative sum of all model responses given by 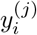.

The objective of the inverse problem is, therefore, to find the vector 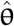 (an estimate of θ) that produces outputs 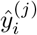 capable of fitting the available observations. The best fit between the responses of the model ŷ ∈ ℝ^*p*^ and the observed data can be estimated in terms of the *residuals*, the difference between observed and predicted measurements, given by r (θ) = ŷ –***𝒟***. The solution to an inverse problem is, in other words, the data fitting whose objective is to calculate an estimate 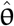 that minimizes some error norm ‖ r ‖ of the residuals [45, 39]. The *least squares* fitting calculates the vector of optimal parameters by taking the mean squared error, given by

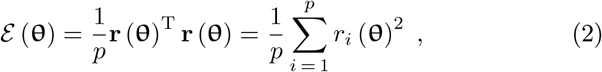

and the estimate 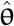 is the vector that minimizes this quantity:

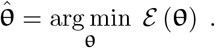

Equation (2) is usually called the *objective function* (or *cost function*). When *ε* → 0, the estimate 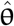 generates an output vector ŷ that has a high level of agreement with the observed data *𝒟*, that is, the residuals are minimized. In general, real problems do not admit *ε*= 0, since the noise that affects the model cannot be predicted with such accuracy.

### 2.4 Bayesian approach for parameter estimation

Bayesian inference provides another perspective for estimating the value of a set of parameters that best characterizes the output of a model, given a set of data. Bayesian inference differs from the deterministic approach because in addition to calibrate the parameter values, it measures their uncertainties, which is one of the focuses of this work. To conduct Bayesian inference, we need some familiarity with a few basic concepts of probability. Here, we give a brief overview of such concepts. A more detailed description is provided by Refs. [47, 22, 3].

Bayes theorem provides a formulation to estimate the posterior probability of the model parameters given a set of observations *𝒟*, based on the likelihood of the event of interest occurring given the prior knowledge on the parameters. The theorem is stated as

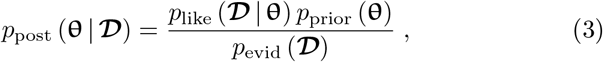

where *p*_like_ (*𝒟* | θ) is the *likelihood* function, *p*_prior_ (θ) is the *prior* information or beliefs on θ, *p*_evid_ (*𝒟*) is the *evidence* related to the observations *𝒟*, and *p*_post_ (θ | *𝒟*) is the *posterior* distribution associated with θ.

A prior knowledge can be thought of as the probability density function over the feasible values of the model parameters, the current knowledge on their values. In turn, the likelihood assumes the role of estimating the probability of characterizing the available data, given a set of parameters. In other words, the likelihood function measures how good the data are being explained by the model. In this work we assume a Gaussian likelihood, which has the form

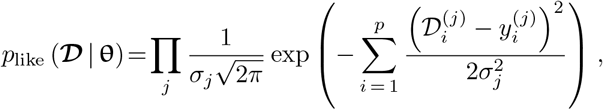

in which 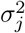 is a measure of the uncertainty (encompassing data errors) related to each quantity *j*, for *j* ∈ {*P, D*}. Here, both 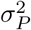 and 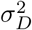 are considered hyper-parameters to be estimated together with the set of parameters θ. The evidence, also referred to as marginal likelihood, is the integral of the likelihood over the prior and is considered as a normalization constant. Thus, we actually evaluate *p*_post_ (θ | *𝒟*) *∝ p*_like_ (*𝒟* | θ) *p*_prior_ (θ) to produce the posterior distribution *p*_post_ (θ | *𝒟*) over the parameters, that is, an updated belief about θ, given *𝒟*.

We conducted the Bayesian inference using the probabilistic programming library PyMC3 [41]. We adopt the Transitional Markov Chain Monte Carlo method proposed by Ching and Chen [8] for parameter estimation (which is not described here, for the sake of brevity).

### 2.5 Gaussian Process Regression

Here we describe the general GP framework, with special attention to regression problems using noisy observations. By definition, a GP is a collection of random variables, any finite number of which have a joint Gaussian distribution [35]. In other words, it is an extension of the multivariate Gaussian distributions to infinite dimensionality. GPRs take place directly in the space of functions, defining priors over functions that, once we have seen some data, can be converted into posteriors over functions [27]. Thus, a GPR model is a Bayesian nonlinear regression model that takes into account the GP prior and whose posterior is the desired regression function that belongs to an infinite dimension random function space [43].

To introduce the GP, denote by t = (*t*_*1*_, …, *t*_*p*_)^T^ the time training points associated to a finite set of *p* observations ***𝒟*** = (*𝒟*_1_, …, *𝒟*_*p*_)^T^. We assume that each observation ***𝒟*** at location *t* is a random variable associated to the GP stochastic process

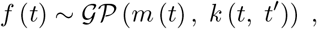

which is completely defined by its *mean function m* (*t*) = 𝔼 [*f* (*t*)], the expected value of all functions in the distribution evaluated for an arbitrary input *t*, and *k* (*t, t*′), the *covariance function* of *f* (*t*), which describes the dependence between the function values for a pair of arbitrary input time points *t* and *t*′, given by *k* (*t, t*′) = 𝔼 [(*f* (*t*) − *m* (*t*)) (*f* (*t*′) − *m* (*t* ′))]. We now consider the regression problem

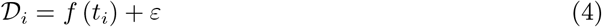

with *ε* being an additive Gaussian *noise* with zero mean and variance *σ*^2^. The GPR begins by assuming the vector-value function **f** ∼ *𝒩* (**0, K**) as the prior distribution, where **K** is the *p × p covariance matrix* whose entries are *k* (*t, t*′). Considering this prior and noise in the time training set, as defined in Eq. (4), the joint distribution taking into account new input time points **t**^*^ and their associated output *𝒟*\* is given by

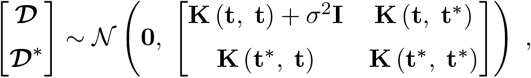

where **I** stands for the *p × p* identity matrix. Therefore, the predictive equations for GPR are derived from the conditional distribution property for the multivariate Gaussian distribution [35]. Considering the Schur complement (for more details on Schur complements, refer to Puntanen and Styan [34]), the posterior predictive distribution is the multivariate Gaussian distribution

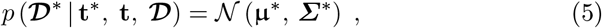

with mean

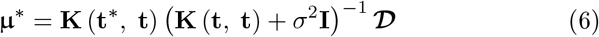

and covariance matrix

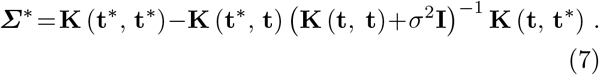

Therefore, the calculation of Eqs. (6) and (7) is sufficient to predict *𝒟*\*. Note that this involves first calculating the four covariance matrices. Furthermore, the covariance depends only on the time training set (**t**) and the new input points (t^*^), and not on the observation measures vector (*𝒟*). By aplying the GPR to the observation set *𝒟*^(*j*)^, for *j* ∈ {*P, D*}, we then obtain 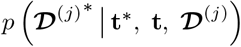, whose corresponding mean values at the time training points are the regularized data used in this work.

The covariance function is commonly called the *kernel* of the GP. This function maps a pair of general input vectors *t, t*′ ∈ **t** into ℝ. The idea behind the kernel is that if *t* and *t*′ are said to be similar, it is expected that the function output (observations) at these points will also be similar. The main attribute of the kernel is to avoid the computation of an explicit nonlinear mapping function that relates input and output data, obtaining the identification of the mapping in the space where the number of parameters to be optimized, the so-called *hyperparameters*, is smaller [20]. Thus, the choice of an appropriate kernel is usually based on prior knowledge related to the behavior of the training data, as for example the occurrence of periodic oscillations, and assumptions such as smoothness [42]. Finding suitable properties for the kernel function is one of the main tasks for defining an appropriate GP.

The kernel can be any function that relates two input vectors, on the assumption that it can be formulated as an inner product, producing a positive semi-definite matrix [5]. Different functions can be combined to produce kernels with a variety of features, generally by both adding and multiplying kernels. Rasmussen and Williams [35] present a comprehensive analysis on the construction of kernel functions. Considering the ability of GPR to approximate the behavior of the used data, we adopt the radial basis function (RBF) *k* (*t, t*′) = *k*_RBF_ (*t, t*′), which is explicitly defined as

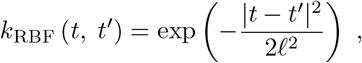

where *𝓁* is the length scale of the kernel.

Rasmussen and Williams [35] present an algorithm for GPR employing Cholesky factorization to solve the matrix inversion required by Eqs. (6) and (7). We use the Scikit-learn library [32] to implement the GPR. Hyperparameters are calculated using a computational routine internal to the library, which uses the L-BFGS-B algorithm [29] to obtain the optimal values. The optimizer is restarted 50 times in order to increase the chances of convergence to the optimal set of hyperparameters. In fact, the number of restarts is an arbitrary choice and, as the GPR is executed only once, the computational cost associated with this procedure is negligible.

### 2.6 Parameter and numerical experiment setups

In models with more compartments and which, in general, have more parameters, finding a unique set of parameter values that best fits some data may be unattainable. Different combinations of parameters can produce similar results for data fitting. This fact is a characteristic of the non-identifiability of parameters [36]. To overcome this issue, some of the biologically-known parameters are fixed, namely the incubation period 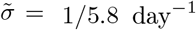 [25], the proportion of symptomatic infected individuals *ρ* = 0.6 [30], and both the recovery rate of infected and positively diagnosed individuals, defined as *γ*_*I*_ = *γ*_*P*_ = 1*/*16.7 day^−1^ [46]. Therefore, the parameters to be estimated are the rate of transmission *β*, rate of removal due to quarantine measures *ω*, and mortality rate *d*_*P*_.

In order to define the initial conditions that make it possible to solve Eq. (1), first consider that the population of the state of Rio de Janeiro is approximately equal to *N* = 17264943 individuals, according to the last demographic census conducted by the Brazilian Institute of Geography and Statistics [6]. Using the reported data, it is possible to define the initial conditions for the number of infected and positively diagnosed individuals, which we assume to be the same at the beginning of the time series. The number of dead individuals on the first day considered in this analysis is also sufficient to define the initial condition for *D*. In addition, it is reasonable to assume *R* (0) = 0, since it is not expected to have recovered individuals at the outbreak of COVID-19. Therefore, the only initial condition for which there is no information from the reported data is related to the exposed individuals. Therefore, we assume *E* (0) as a parameter to be estimated and the initial condition for the number of susceptible individuals is given by *S* (0) = *N–* (*E* (0) + *I* (0) + *P* (0) + *R* (0) + *D* (0)).

We partition data sets into two subsets, which we call training data and test data. The key idea behind this approach is to analyze the predictive capacity of the model, by comparing test data with short-term simulations, which are calculated using parameters estimated with training data. In this way, it is possible to compare the gain of using regularized data in the parameter estimation procedure, analyzing the results considering an actual scenario. Furthermore, we only consider shortterm predictions in our analyzes since, as the simulations are compared with original data. Therefore, we analyze scenarios in which we adopt 14 data points in the test set. Of note, other time windows could also be used.

Arbitrarily, we choose the minimum size of the training data set equal to 60. We set up 14 values in the test data set and calculate successive estimates of the parameters of the model, gradually increasing the proportion between training and test data. After each run, new data is added to the training set, so that the test set is composed of the next 14 values in the time series. As data for 210 days are available, 136 sets of parameters are estimated, using the deterministic approach described in Section 2.3. This procedure is performed both using training data as it stands, and after regularization. The simulations using the optimal parameters are compared to the corresponding test set (without being regularized), by the computation of the normalized root-mean-square error (NRMSE) [15], considering both the cumulative number of infected and dead individuals—in this step, the cumulative data is adopted, with the purpose of minimizing the influence of noise. The root-mean-square error is normalized by the difference between the highest and lowest values in the corresponding data set.

All optimal parameters are calculated by combining the Differential Evolution [44] and the Nelder-Mead Simplex [28] methods. For each problem, the solution is estimated by Nelder-Mead Simplex and refined by Differential Evolution, which searches for the optimal parameters in the vicinity of the previously obtained point. The solution to each problem—the best individual in the population—is taken as an initial estimate for the next problem. Nelder-Mead Simplex runs with coefficients of reflection, expansion, contraction, and shrinkage equal to 1, 2, 0.5, and 0.5, respectively (as commonly adopted in the literature). In turn, Differential Evolution runs with 20 individuals in the population, amplification factor equal to 0.6, and crossover probability equal to 0.95. The search space is bounded by 0 ≤ *β* ≤ 10^−6^, 0 ≤ *ω* ≤ 1, 0 ≤ *d*_*P*_ ≤ 1, and 0 ≤ *E* (0) ≤ 10^4^ in appropriate units (a. u.).

## 3 Results and Discussion

### 3.1 Influence of regularization of data

Initially, the data used in this analysis is regularized using GPR as described in Section 2.5. The values of the hyperparameters tunned for the RBF kernel are *𝓁* = 68.9 and *𝓁* = 50.2 for daily data of infected and dead individuals, respectively. Figure 2 shows an illustrative comparison between the original data for daily infected and dead individuals and the corresponding regularized data. Shaded areas represent two standard deviations from the fitting data. We also show the respective cumulative data, in order to allow a visual inspection of the agreement of the data resulting from the regularization, but smoothing out the noise.

**Fig. 2.**
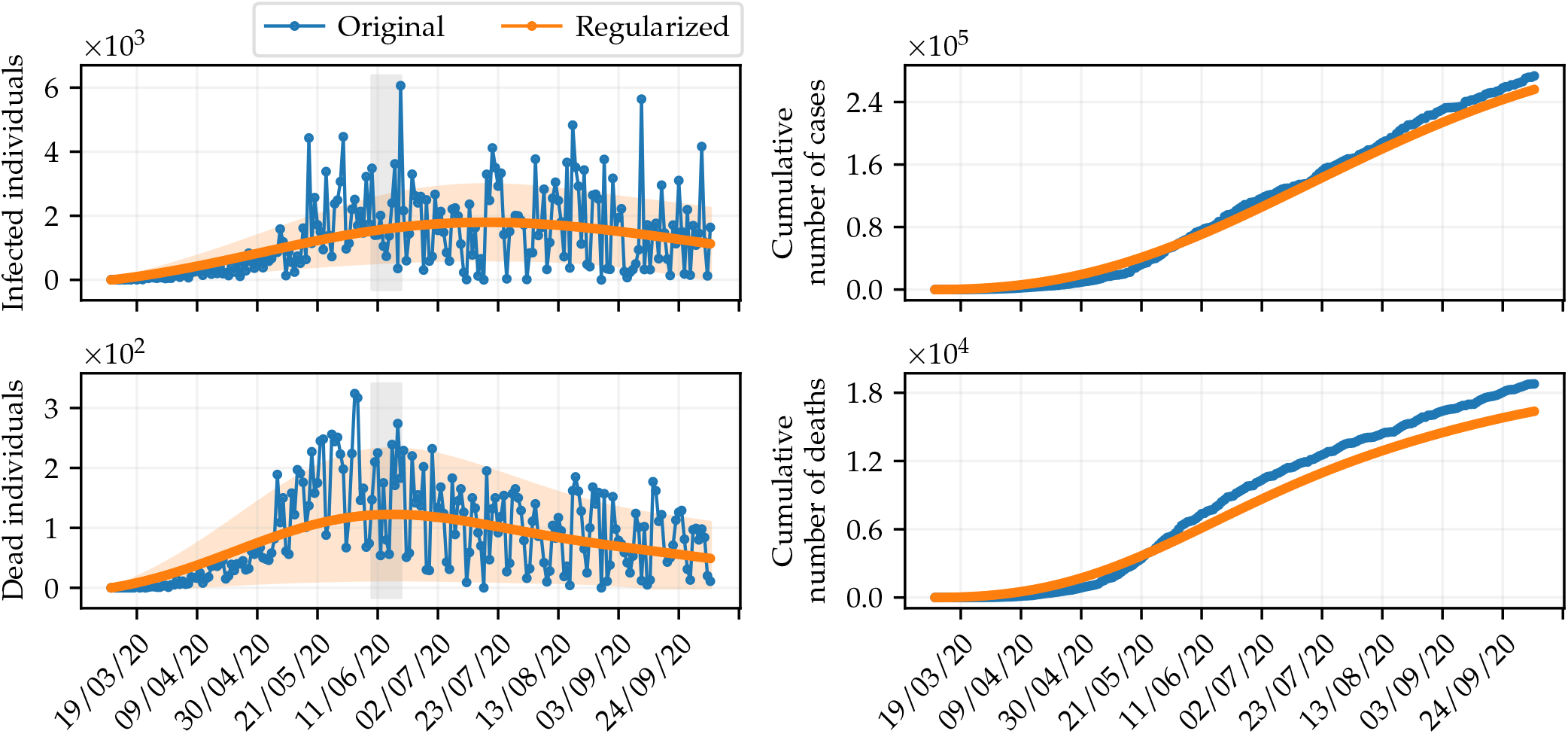
Results of the regularization of data using GPR. The blue dots are the original data and the orange dots represent the resulting regularized data. Shaded areas indicate two standard deviations from the corresponding regularized data. We also show the cumulative data, for comparison purpose.

Next, the influence of estimating the parameters of the SEIRPD-Q model, presented in Section 2.1, using the regularized data set, in relation to the approach that adopts the original data is analyzed. Our focus is to assess the gain related to the regularization of data within the scope of compartmental models and, therefore, we consider the simulations using the model adopted in this analysis. It is worth mentioning that the proposed analysis can be extended to any compartmental model with a structure similar to the model given by Eq. (1). The analysis is performed by evaluating the NRMSE between model predictions and test data.

Results concerning the optimal parameters calculated by Differential Evolution and Nelder-Mead Simplex (Section 2.6) are shown in Fig. 3. For each set of parameters obtained in the 136 runs using the methodology described in Section 2.6, considering each type of data (original and regularized), the model is simulated and we calculate the corresponding NRMSE. Then we compute the area under the curve, which is composed of the values of the NRMSEs. Analyzing Fig. 3, the effect of data regularization on parameter estimation is clear: for most of the estimated parameter sets, the corresponding simulations have better agreement with the test data. Translating into numbers, regularized data resulted in more reliable predictions in 64.71% of runs. The area under the curve for original data is approximately equal to 899.66 u. a. (unit of area), whereas for regularized data it is 705.70 u. a. This represents an average improvement of approximately 21.56%. Note that the effect of the regularization is more prominent when the data set to be fitted is larger, since the influence of the noise tends to become more intensive.

**Fig. 3.**
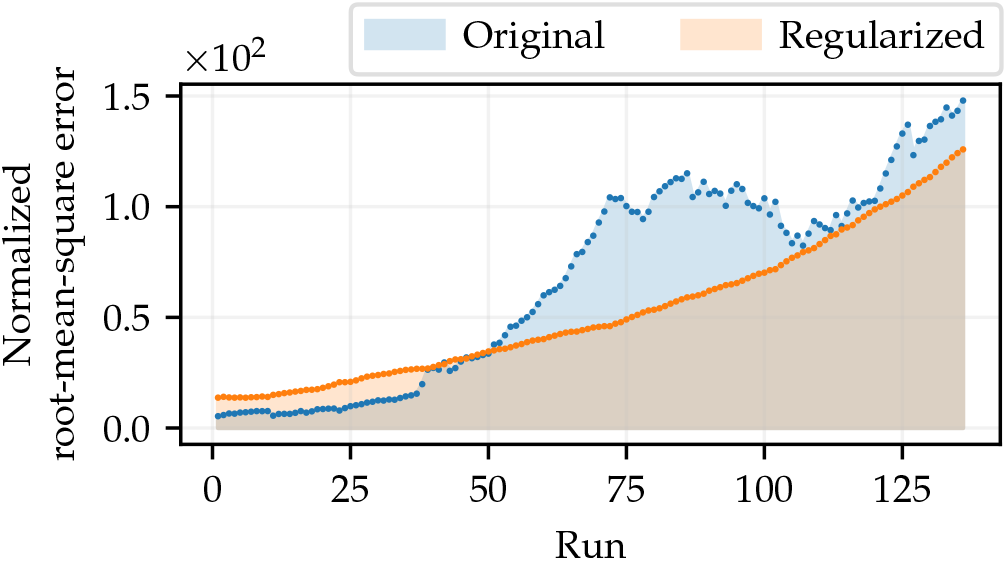
NRMSEs computed by comparing simulations of the SEIRPD-Q model, performed using parameters that best fit the training data, where θ = (*β, ω, d*_*P*_, *E* (0)), in relation to the test set (composed of 14 points). The area under the curve represents the total deviation relative to the test data in all runs, where the number of training data varies.

The implication of using regularized data is even more straightforward when we analyze the variability of simulations resulting from the optimal parameters corresponding to each point in Fig. 3. For this purpose, consider the results shown in Fig. 4, in which the shaded area represents the range of the simulations related to the daily number of infected and dead individuals, for both original and regularized data, whose parameters are estimated by varying the amount of training data, as previously described. In turn, the points represent the corresponding data, which together with the box-and-whisker diagrams, aim to demonstrate the variability of the simulations on specific days. We also show the corresponding cumulative values of the results obtained, in order to provide better conditions for comparison. It is worth mentioning that the set of training data, even presenting variable size considering each set of parameters that is estimated, is completely shown in all cases.

**Fig. 4.**
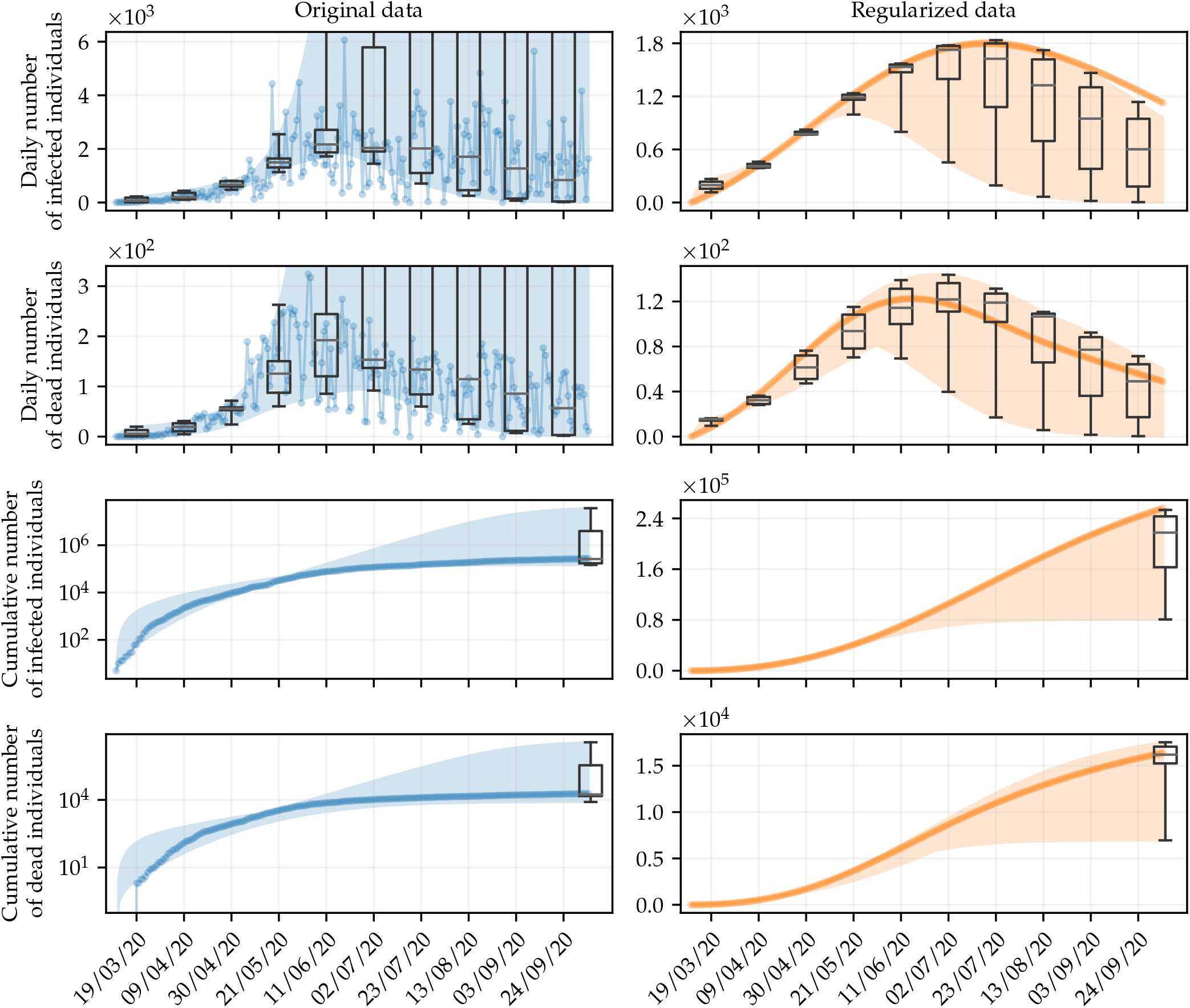
Simulations of the daily number of infected and dead individuals using optimal parameters obtained with the deterministic approach, by varying the amount of data in the training set. The shaded areas represent the variation range of the simulations, whereas the points are the fitted data. Box-and-whisker diagrams are used to show the variability of the simulations on specific days. We also show the corresponding results for cumulative data, where the noise is less effective, for comparison purposes. The results follow the same color scheme as in Fig. 2, blue for original data, and orange for regularized data.

It is clear that simulations resulting from parameters estimated using data with no regularization have more variability than after regularization. Considering that on October 5, 2020 (the last day of the simulation), Rio de Janeiro had 273,335 confirmed cases, the boundary values of the shaded area on this day, for the cumulative number of infected individuals obtained with original data are 146,763.4 and 36,518,946.5, whereas the quantities obtained with regularized data vary between 80,762.4 and 253,387.8. In the case of dead individuals, such values vary between 8,066.9 and 3,579,976.6 considering original data, and between 6,950.6 and 17,528.7 considering regularized data, with the actual cumulative number of dead individuals on this day being 18,780.

The dispersion of these results can be alternatively analyzed using a box-and-whisker diagram. The boxes are bounded by the lower and upper quartiles (which we call Q1 and Q3, respectively) of the full model prediction values, and the horizontal line inside the box represents the median. Whiskers, the vertical lines bounded by perpendicular dashes, extend from the minimum value of the data to the first quartile, and from the third quartile to the maximum value of the data. Whiskers express the variability outside these quartiles. They range from the lowest to the highest values of each set, both for original and regularized data, since no simulation can be considered an outlier. The statistical results associated with the box-and-whisker diagrams in Fig. 4 are summarized in Table 1. It is clear that regularization using GPR preserves the behavior of the data, although substantially reducing the variability of the simulations. The reduction in variability provides evidence that regularizing the data with the GPR does not cause problems related to overfitting. Simulations influenced by overfitting would deviate significantly from the real data. In this case, parameter estimation using regularized data provides a means of reducing noise without negatively influencing forecasts [26].

**Table 1.** Statistical results of all 136 simulations performed using the optimal parameters. The results refer to the cumulative number of infected and dead individuals, that is, 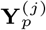 for *j* ∈ {*P, D*}, obtained using both original and regularized data on the last day that the model was simulated, October 5, 2020. On this day, Rio de Janeiro accumulated 273,335 confirmed cases and 18,780 deaths.

|  | Original data |  | Regularized data |  |
| --- | --- | --- | --- | --- |
| | $\mathbf{Y}_p^{(P)}$ | $\mathbf{Y}_p^{(D)}$ | $\mathbf{Y}_p^{(P)}$ | $\mathbf{Y}_p^{(D)}$ |
| Min | 146,763.4 | 8,066.9 | 80,762.4 | 6,950.6 |
| Max | 36,518,946.5 | 3,579,976.6 | 253,387.8 | 17,528.7 |
| Median | 259,406.5 | 17,313.5 | 217,621.5 | 16,196.7 |
| Q1 | 172,065.9 | 14,796.5 | 163,064.3 | 15,249.4 |
| Q3 | 3,970,094.4 | 342,994.5 | 243,274.8 | 17,059.5 |

Now consider a forecast of the epidemic in terms of the cumulative number of infected and dead individuals, aiming to show the difference of the simulations in relation to the analyzed data. For this purpose, we adopt the Bayesian approach for parameter estimation, presented in Section 2.4. We arbitrarily choose the training data sets for the daily number of infected and dead individuals, both original and regularized, with 196 values, the maximum number of elements that the test set can contain. In turn, the test set is made up of the next 14 values in the time series. By this analysis, we present a visual perspective of the benefit of using regularized data in the parameter estimation procedure, providing a way of comparing the agreement between simulations and test data.

The parameters to be estimated are the same as in the previous analysis, and are assumed to be uniformly distributed, in such a way that *β* ∼ *𝒰* (0, 10)^−6^, *ω* ∼ *𝒰* (0, 1), *d*_*P*_ ∼ *𝒰* (0, 1), and *E* (0) ∼ *𝒰* (0, 10^4^) in a. u. Table 2 shows the *maximum a posterior* (MAP) estimates of the parameters, as well as the corresponding 95% credible interval (CI) [3]. Likewise, all other parameters of the SEIRPD-Q model are taken as biological parameters and have the same values previously reported (see Section 2.6). Figure 5 shows the posterior distribution of the estimated parameters of the SEIRPD-Q model, obtained when original (blue bins) and regularized (orange bins) data are employed for calibration. In the right frame, violin plots express the variance of the inferred parameters using original and regularized data. Of note, the latter is much narrower than the former. This fact is reflected in the stochastic simulation of the SEIRPD-Q model shown in Fig. 6, along with the training and test data sets (note that the latter is never regularized). The solid curves represent the model responses when the free parameters are set to be the respective MAP values, shown in Table 2. In turn, the shading around the curves represents the 95% CI, the discrete points are the available data, and the ranges of the training and test sets are colored in gray for daily and cumulative data, respectively. Results related to infected individuals are shown in red, whereas dead individuals are shown in green.

**Table 2.** MAP values and 95% CIs of the parameters estimated using Bayesian calibration (in a. u.).

|  | Data type |  |
| --- | --- | --- |
|  | Original | Regularized |
| $\beta$ | $1.5248 \times 10^{-8}$ | $1.2419 \times 10^{-8}$ |
| | $(1.2795, 1.7991) \times 10^{-8}$ | $(1.2206, 1.2639) \times 10^{-8}$ |
| $\omega$ | 0.007121 | 0.004859 |
|  | (0.005704, 0.008576) | (0.004738, 0.004996) |
| $d_P$ | 0.07126 | 0.0625 |
|  | (0.06215, 0.08161) | (0.06030, 0.06475) |
| $E(0)$ | 1190.8021 | 1236.5740 |
|  | (655.0367, 1818.3565) | (1168.6603, 1305.0816) |

**Fig. 5.**
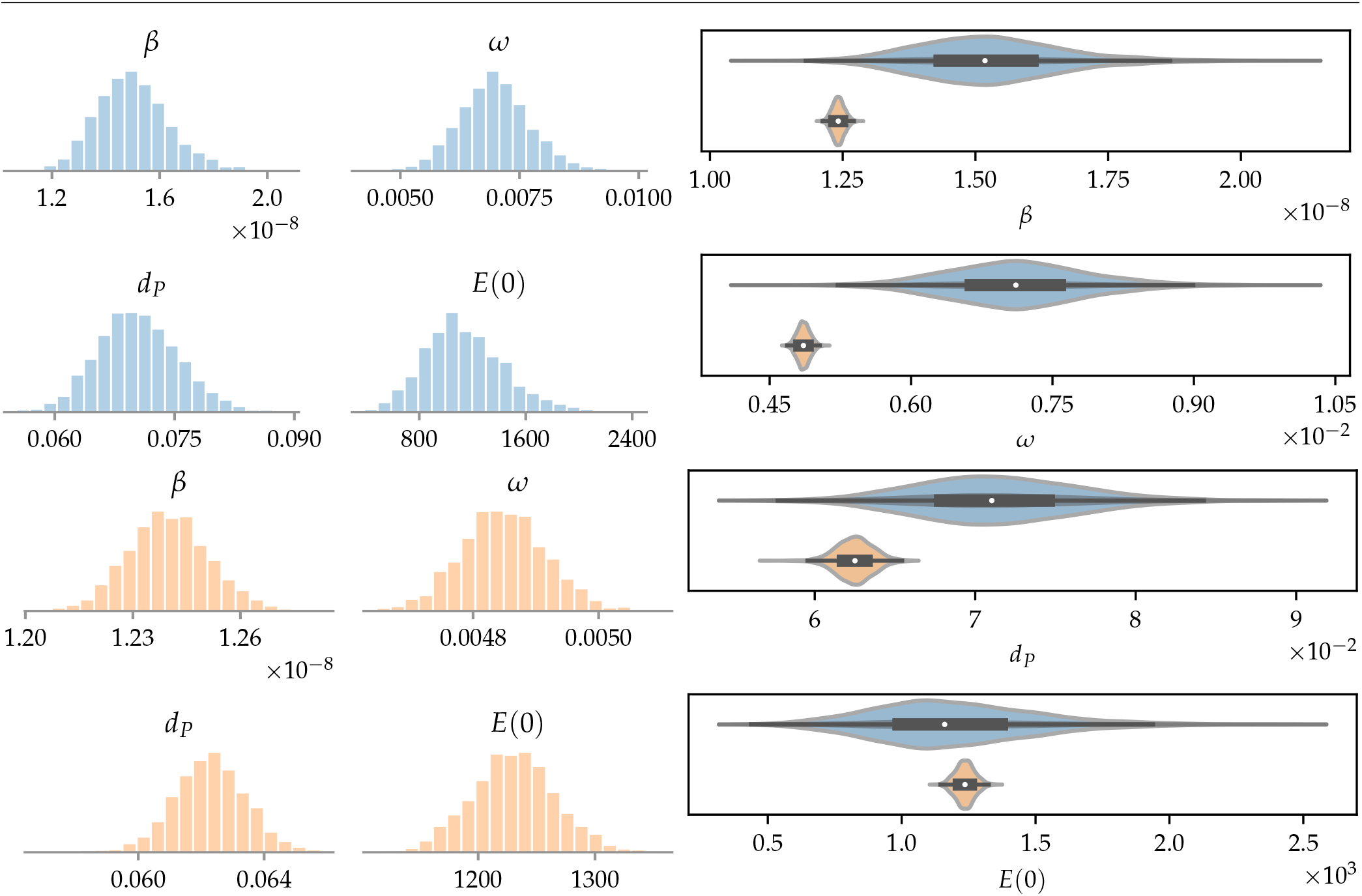
The left frame shows the posterior distribution of the parameters obtained in the Bayesian inference for original (in blue) and regularized (in orange) data, by fitting the daily number of infected and dead individuals; the right frame illustrates the variance of each parameter in a comparative way, where the same color scheme is adopted.

**Fig. 6.**
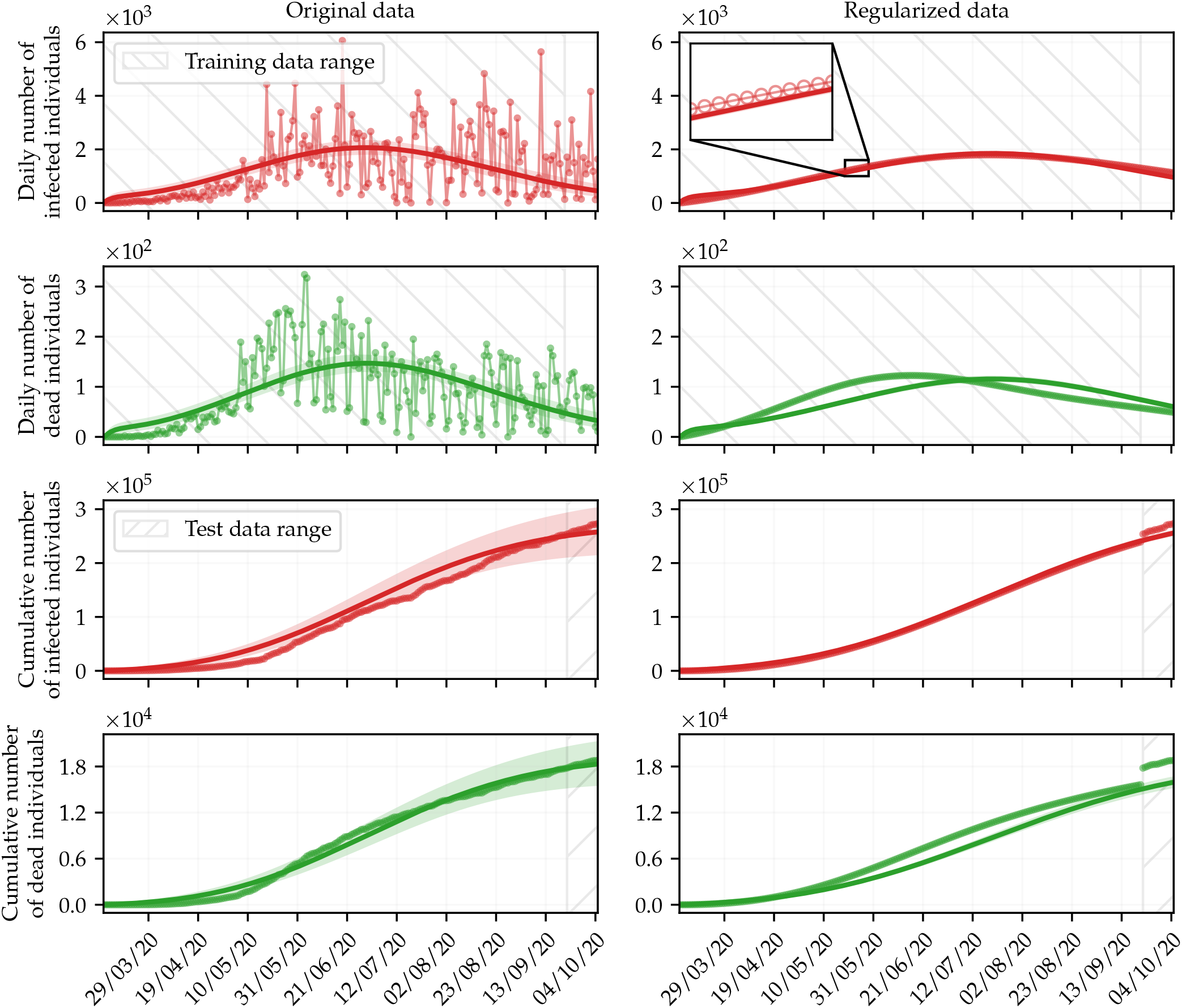
Simulations with parameters obtained by fitting the daily number of infected (in red) and dead (in green) individuals using the Bayesian approach. The shaded areas refer to the 95% CI (see Table 2). Hatched areas bound the range of training and test data. Training data is never regularized.

Analyzing Fig. 6, it is clear that the Bayesian inference obtained suitable results, using both original and regularized data. The deviation between the simulations and the cumulative data in the test data range when regularized data are used for calibration is due to the regularization calculated using GPR, as can be seen in Fig. 2. Nevertheless, the regularization of the training data seems to make the predictive capacity of the model less unstable. Thus, the model could better capture the behavior of the data, eventually leading to more appropriate predictions. Of note, the narrower posterior distributions obtained by performing the calibration with the regularized data resulted in less uncertainty about the values predicted by the model, as can be seen in Fig. 6. This is because the posterior distributions obtained when data are regularized tend to have a smaller standard deviation, as can be seen in the right frame of Fig. 5 and in the CIs shown in Table 2.

### 3.2 Influence of time-varying parameters

A further feature that may improve the predictive capacity of compartmental models is the adoption of time-varying parameters. In an effort to analyze this aspect, consider the 136 sets of optimal parameters, obtained using the deterministic approach that led to the results shown in Fig. 3. Figure 7 shows the values of the calibrated parameters for each corresponding run, both using original and regularized data. Initially, note that the variability of the set of parameters obtained by fitting the original data is much higher than those referring to regularized data. This is a consequence of the high level of noise in the data. Note, for instance, the gray shaded areas in Fig. 7. They refer to the same runs for all parameters and correspond to the shaded area in the same color in Fig. 2. In this time window, the data show very large variations on subsequent days, especially those of the daily number of infected individuals. These variations influence the values of the parameters obtained, giving rise to a great difference in the value of the set of parameters that best fits the data just by adding a new single value to the training set, which does not occur with regularized data.

**Fig. 7.**
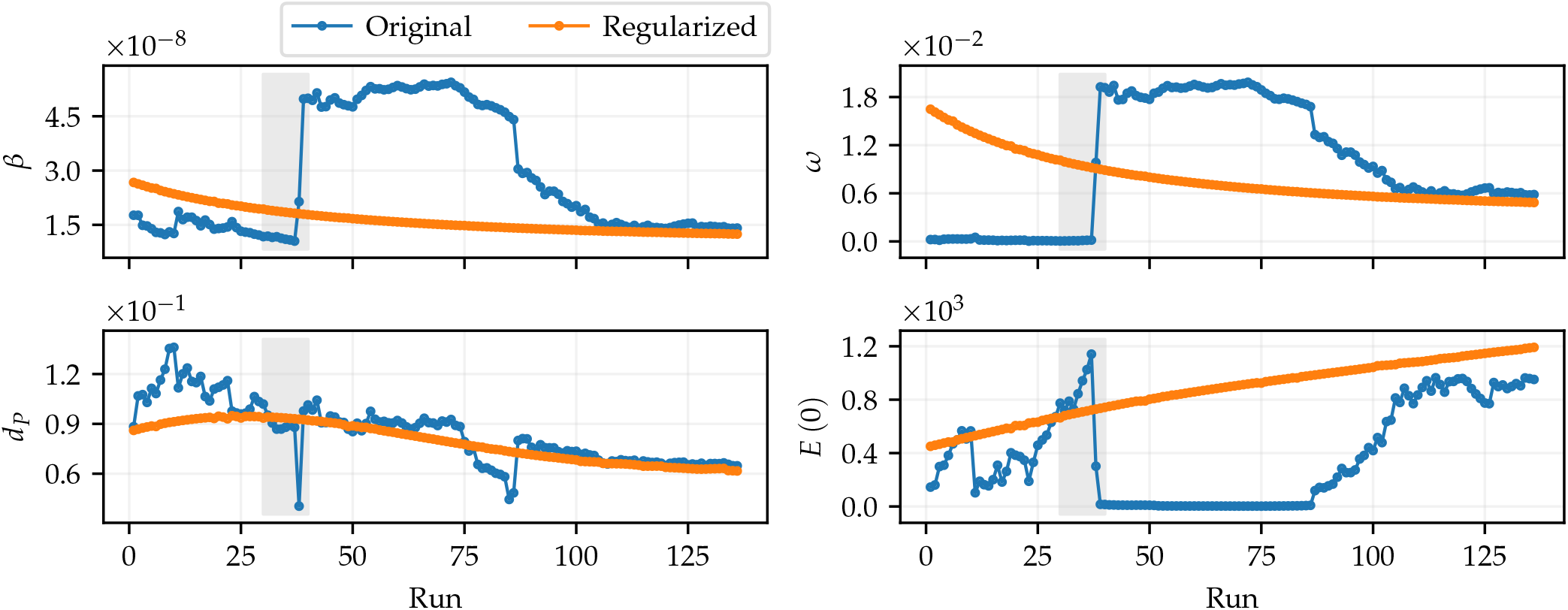
Optimal parameters obtained by fitting the daily data of infected and dead individuals, for θ = (*β, ω, d*_*P*_, *E*(0)). Each run is associated with a training set with a specific size, from 60 to 196 data in each set. The test data refer to the 14 subsequent data from the corresponding run. Parameters obtained by fitting original data are shown in blue, and with regularized data are shown in orange.

The optimal parameters in Fig. 7 express some meaningful facts: first, the behavior of the initial conditions for exposed individuals reveals that defining this value just as a fixed proportion of the population size can undermine the capacity of the method for solving the system of differential equations; second, the parameters *β* and *ω* exhibit similar behaviors (especially when considering the results obtained with regularized data). Over time, the rate of contact between susceptible and infected individuals decreases (assuming the hypothesis that recovered individuals are immune for some time), so that quarantine measures end up being eased, which is reflected in the reduction of isolation measures. Definitely, political and social interests also play a significant role in this behavior.

The inspection of the behavior of *d*_*P*_ in Fig. 7 indicates that the mortality rate of positively diagnosed individuals basically only decreases after a certain run, around the gray shaded area. This behavior suggests that the parameter can be approximated by a function. In this case, we propose to represent *d*_*P*_ as a function of the form

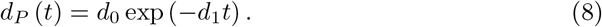

This approach assumes an inherent error related to the first runs, at expense of better representing the parameter behavior in later times. Thus, *d*_*P*_ becomes variable over time, and the vector of parameters to be estimated is formed by θ = (*β, ω, d*_0_, *d*_1_, *E* (0)).

Consider the methodology previously adopted, where the set of training data is gradually expanded with each parameter estimation, but now taking into account the proposed approximation for *d*_*P*_ (*t*) defined in Eq. (8). The search intervals for the new parameters are 0 ≤ *d*_0_ ≤ 1 and 0 ≤ *d*_1_ ≤ 1 and all other parameters follow the same specifications defined before. Figure 8 shows that this strategy is reflected in the reduction of the NRMSE considering the test data set with 14 values, in most simulated forecasts. The NRMSEs considering regularized data and time-varying *d*_*P*_ (orange dots) represent a better approximation of the test data set in 73.53% of the analyzed runs, in relation to the results considering original data (blue dots). The area under the curve obtained with regularized data and *d*_*P*_ (*t*) is 733.06 u. a., whereas for original data the area is equal to 1427.28 u. a., which represents a reduction of approximately 48.64%.

**Fig. 8.**
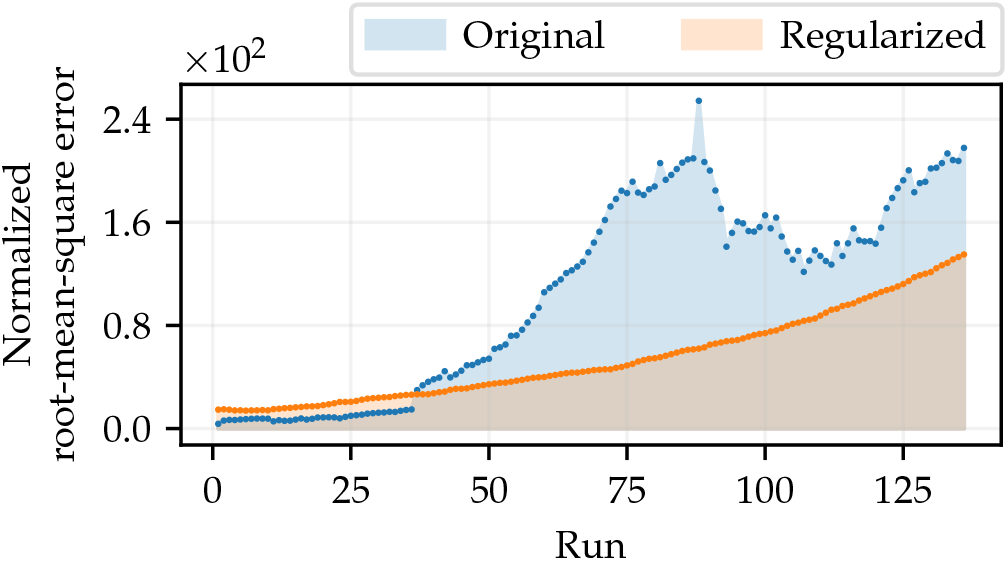
NRMSEs computed by comparing simulations of the SEIRPD-Q model, performed using parameters that best fit the training data, where θ = (*β, ω, d*_0_, *d*_1_, *E*(0)), in relation to the test set (composed of 14 points). The area under the curve represents the total deviation relative to the test data in all runs, where the number of training data varies.

As in Fig. 7, we are interested in understanding the behavior of the parameters inherent to the function when *d*_*P*_ varies over time. Figure 9 shows the parameters *d*_0_ and *d*_1_, as well as the other calibrated parameters of the model, calculated in each parameter estimation procedure using both original and regularized data. The color scheme follows what has been adopted, blue for original data and orange for regularized data. In addition, Fig. 9 shows the range of Eq. (8), taking the optimal values of *d*_0_ and *d*_1_, alongside the values of *d*_*P*_ (shown in Fig. 7) for both original and regularized data. In this regard, we are interested in analyzing the behavior of *d*_*P*_ (*t*) in terms of the values obtained for the case where *d*_*P*_ is constant.

**Fig. 9.**
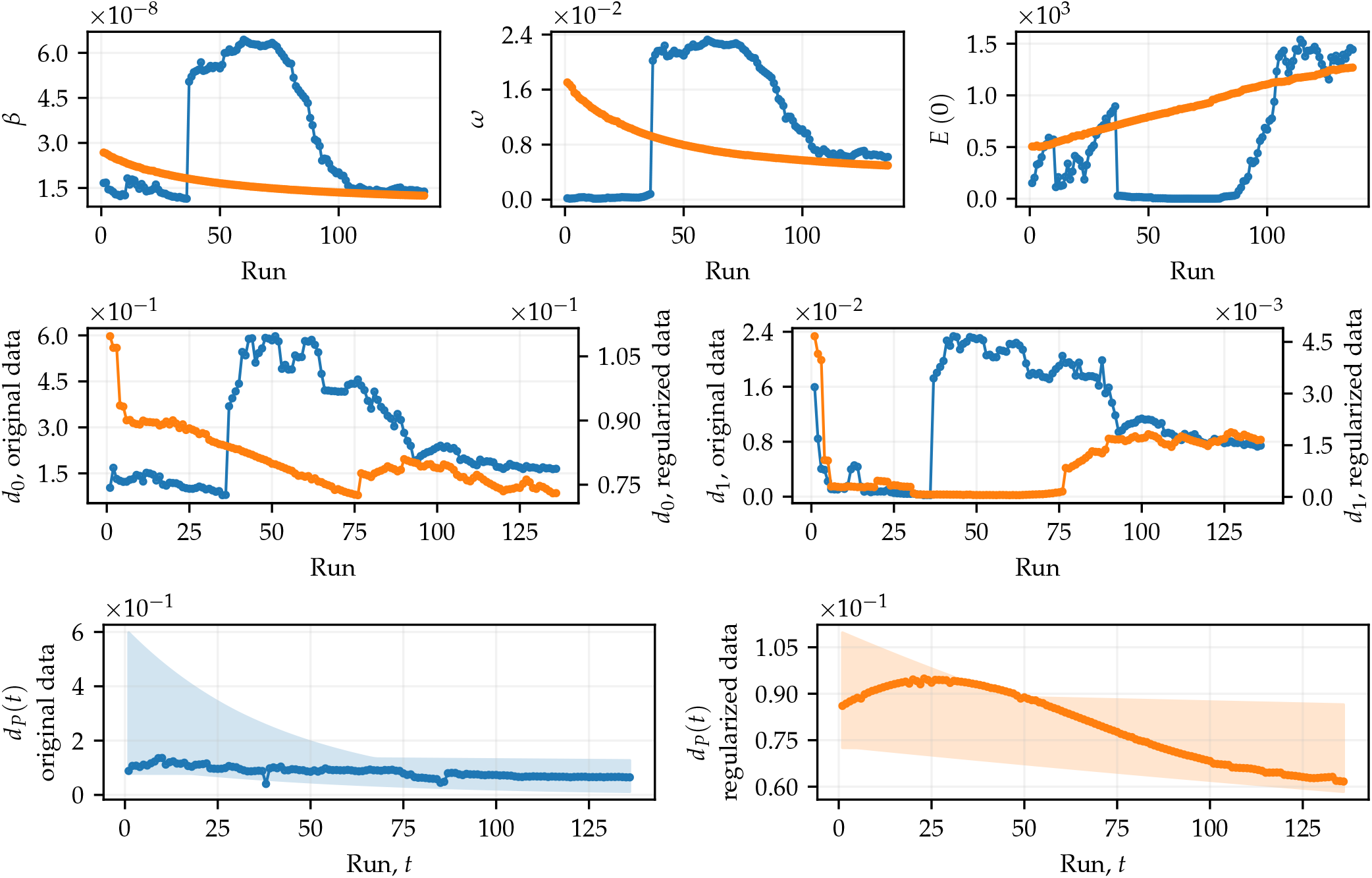
Optimal parameters obtained in a procedure similar to that of Fig. 7, for θ = (*β, ω, d*_0_, *d*_1_, *E*(0)). We also show the interval that includes the curves of *d*_*P*_ (*t*), for each optimal value of *d*_0_ and *d*_1_ associated with the same run. In addition, we also show the optimal values of *d*_*P*_ presented in Fig. 7, for purposes of comparison with the curves of *d*_*P*_ (*t*).

In the first runs, *d*_*P*_ (*t*) is expressed by a nearly constant function, since the values of *d*_1_ are very close to zero (see the behavior of *d*_1_ in Fig. 9). In this case, *d*_*P*_ (*t*) ≈ *d*_0_. These curves practically coincide with the first points shown in the frame corresponding to *d*_*P*_ (*t*) obtained with regularized data in Fig. 9, which means that, in fact, the calibrations are quite similar. This behavior occurs because, given the training set for such runs, the model identifies that the mortality rate is not decreasing and, therefore, the calibration procedure estimates the most suitable function for such data (by means of *d*_0_ and *d*_1_), which in this case is nearly constant, without loss of generality.

As the training set gets larger, *d*_*P*_ (*t*) starts to behave as expected, exponentially decreasing. In this case, the first calibrations provide functions relatively distant from the corresponding points in Fig. 9, especially in the early times. In the last runs, the functions show good agreement with the compared points. However, it is important to note that *d*_*P*_ (*t*) is not expected to represent the exact behavior of such data in the long run. In general, when the mortality rate varies over time, the compartmental model may be more capable of capturing the dynamics of the data, allowing for more accurate predictions. This hypothesis is supported by the last results obtained in Fig. 8, where the orange dots always represent the best approximation in relation to the test data.

## 4 Conclusions

Our study provides a framework that aims to increase the predictive capacity of compartmental models. This is relevant from the point of view that the proposed strategies can be extended to other data sets and compartmental models. Especially in the context of the epidemiological modeling of COVID-19, approaches of this type can be useful, taking into account the wide range of existing compartmental models and the fact that the data analyzed here are similar to others regarding noise. This study has gone some way towards enhancing our understanding of the influence of noise on the estimation of parameters of compartmental models. The work has revealed that the regularization of data by means of GPR can represent an alternative to mitigate the effect of noise in a given parameter calibration. Since this procedure must not be repeatedly applied, in the context to which it is proposed, the computational cost of the GPR can be considered irrelevant.

Our research also suggests that it may be useful to use time-varying parameters, to the detriment of the usual approach that adopts constant parameters. It incorporates additional degrees of freedom into the model, in order to provide more flexibility to describe the behavior of the analyzed data. The choice of such a function depends on several factors, as for example the physical meaning of the parameter and the additional parameters inherent to the function. This analysis can be conducted for any model and, clearly, the gain is related to the appropriate choice of the function, especially if many parameters to be estimated are included.

## Data Availability

The source code used to generate the results is publicly available at https://github.com/gustavolibotte/enhancing-forecast-COVID-19.

## Availability of data and material

The data used in this work are publicly available according to Ref. [10].

## Code availability

The source code used to generate the results is publicly available at github.com/gustavolibotte/enhancing-forecast-COVID-19.

